# Sleep Intervention for NHS Healthcare Shift Workers: A Pilot Study of Noise-Masking Earbuds

**DOI:** 10.64898/2026.08.28.26361673

**Authors:** Robert Hickman, Dan W. Joyce, Nicholas Gray, Sukhi Shergill, Teresa C. D’Oliveira

## Abstract

**Background:** Shiftwork disrupts natural sleep-wake cycles, alters light exposure patterns, and contributes to circadian misalignment. Detrimental health consequences associated with shift work include elevated risk for metabolic disorders, cardiovascular disease, cancer and all-cause mortality. Healthcare workers have one of the highest rates of shift work exposure, yet there are relatively few non-pharmacological interventions (with good evidence) developed to improve sleep outcomes in this population.

**Objective:** A pre-post pilot interventional study assessed the acceptability and perceived effectiveness of commercial noise-masking earbuds on improving subjective sleep characteristics among National Health Service (NHS) healthcare staff working fast rotating shifts.

**Methods:** Noise-masking sleep earbuds (Kokoon NightBuds) were worn for a pilot six-week intervention by twenty-seven NHS nurses (aged 26-43 years, 88.9% female) working fast rotating shifts from the *EClocker Study*. Sensors inside the earbuds were paired with a smartphone app to monitor sleep. An audio library in the smartphone app delivered personalised relaxation exercises and sleep techniques drawn from cognitive behavioural therapy for insomnia (CBT-I). A pre-post two-week monitoring period with daily smartphone-based Experience Sampling Methods (ESM) captured perceived daily sleep patterns. Acceptability and perceived effectiveness of the earbuds in promoting better sleep outcomes was assessed.

**Results:** Use of the noise-masking sleep earbuds over a six-week period was associated with positive sleep improvement trends and elicited promising acceptability. Almost two thirds of NHS fast rotating shift nurses (63%) subjectively reported reductions in general sleep disturbance symptoms (PSQI Global), one in four experienced perceived sleep quality improvements (SQ; 25.9%), one in five reported sleeping longer (TST; 22.2%), and a third perceived falling asleep faster (SOL; 33.3%), had better sleep efficiency (SE; 33.3%) and improved daytime dysfunction (33.3%) (PSQI subcomponent scores). Sleep diaries (CSD) collected daily using smartphone-based ESM also demonstrated small improvements post-sleep earbud use; nurses reported sleeping an average 18 minutes longer (TST) and fell asleep more easily, on average 11 minutes faster (SOL). Sleep earbuds were generally well tolerated; 56% of nurses reported the earbuds as (somewhat to very) helpful, 52% reported (somewhat to strongly) falling asleep more easily (SOL), 44% felt (somewhat to strongly) their sleep quality was improved (SQ) and 30% agreed (somewhat to strongly) they slept longer (TST) and had less disturbed sleep.

**Conclusions:** To our knowledge, this is the first study in Europe to pilot noise-masking earbuds as a potential non-pharmacological aid to improve sleep-wake behaviours or mitigate fatigue for healthcare staff. Preliminary results showed promising acceptability and (small) perceived sleep improvement trends following a targeted six-week earbud intervention in NHS fast rotating shift nurses.

## Introduction

### Background

Although most shift working healthcare staff express strong interest in undertaking sleep-targeted interventions [1–3], there are limited high quality and well-tested research studies investigating their comparative effectiveness in promoting better sleep health for nurses [4, 5]. Evidence for which specific treatment or intervention may work best for a particular shift work individual is also lacking. Existing interventions have traditionally been generalised, focused at the organisational or broad institutional level (e.g., adjusting shift schedules, working time arrangements, or forward rotating roster systems) [6–12] and there are limited ‘multi-component’, individualised approaches for shift workers [13–16].

Few studies have evaluated non-pharmacological, ‘individually-directed’ interventions to improve sleep in NHS shift workers [14]. Among nurses, these include promoting sleep hygiene strategies or sleep health awareness [6, 17–19], fatigue countermeasures such as prophylactic naps [20–24], physical activity or diet [25, 26], behavioural approaches such as tailored CBT-I [27–30], mindfulness [31, 32], light exposure or light therapy [12, 33–38], music therapy and white noise [39] and melatonin after nightshifts [40]. An ongoing non-randomised trial (OxBIS; NIHR203667) is also evaluating a behavioural intervention for NHS shift workers. Digital mobile health app interventions have emerged to improve sleep among shift workers [41–44] or to treat sleep-wake disorders [45] such as *Sleepio* (*SleepioRx*; FDA-cleared in the USA), which is clinically- evidenced for insomnia symptoms and the first digital therapeutic to receive National Institute for Health and Care Excellence (NICE) guidance [46, 47]. A recent study trial with a sample of shift working nurses also had positive results for a mobile app (*SleepSync*) providing personalised sleep-wake and behavioural recommendations tailored for shift workers [48]. Alternative mobile apps include *Timeshifter* or *Arcashift* to aid shift workers, *Chronoshift*, *StopJetLag* or *Uplift* to prevent jet lag, and more general sleep tracking apps such as *RISE Science* [49]. Many of these digital mobile apps, particularly for shift workers, lack peer-review and evidence-based studies that validate their efficacy [42, 50].

Night shift nurses sleeping in the daytime also report additional environmental challenges related to noise disturbances [51]. A systematic review and meta-analysis of 38 studies found that continuous white noise at night may aid sleep by masking disruptive sounds or providing monotonous stimulation [52]. However, the evidence for white noise is not well-established and exposure to continuous sound may even interrupt or compromise sleep processes (e.g., perturb slow wave or REM sleep) [52].

Three recent studies evaluated the Bose noise-masking SleepBuds (Bose Corporation, Framingham, MA, USA) to foster improved sleep outcomes among day shift [53] and night shift [54] healthcare workers and healthy volunteers who had experienced nightly environmental noise disturbance [55]. To our knowledge, no research to date has evaluated similar commercial sleep earbuds in Europe or with UK healthcare staff. Additionally, a recent scoping review [4] and systematic review [5] reported no intervention studies for nurses conducted in the UK designed to promote sleep or ameliorate detrimental fatigue. This was the primary objective of *EClocker Phase 2* and aimed to address the paucity of accessible and individualised sleep interventions for UK healthcare workers. In *EClocker Phase 2*, commercially available sleep earbuds (*Kokoon NightBuds*; Kokoon Technology Ltd.) were piloted with NHS nurses working fast rotating shifts from *EClocker Phase 1*. These earbuds contain sensors that can fade audio and introduce either coloured noise to mask disturbances or switch off the device – as identified by Riedy, Smith [52], auto-fade noise-masking may aid sleep onset but prevent continuous stimulation and activity of the auditory system overnight.

### Objectives

The purpose of the *EClocker Phase 2 Study* was twofold. Firstly, to explore nurses’ perceptions, acceptability, and the initial implementation of a 6-week pilot sleep intervention (noise-masking earbuds) for a cohort of NHS fast rotating shift nurses. Secondly, to report on preliminary data on sleep changes from before and after the pilot sleep intervention. Any potential sleep improvements during the pilot study were expected, in-turn, to have downstream benefits on NHS nurses’ affective states, emotional reactivity, and reduced burnout.

## Methods

### Participants and Study Design

NHS staff were fully qualified and registered NHS nurses with the Nursing and Midwifery Council (NMC), working at three London NHS Foundation Trusts. Thirty-three fast rotating shift nurses from *EClocker Study Phase 1* enrolled in a pilot (non-randomised) sleep earbud intervention [design schematically shown in **Figure 1**] and has previously been described elsewhere. Six nurses subsequently withdrew from the study (unexpected personal reasons or non-response), resulting in a dropout rate of 18%. A final sample of twenty-seven NHS nurses completed the *EClocker Phase 2* sleep intervention (82% completion rate). The sample size was comparable to prior research using noise-masking earbuds in healthcare shift workers [54].

**Figure 1.**
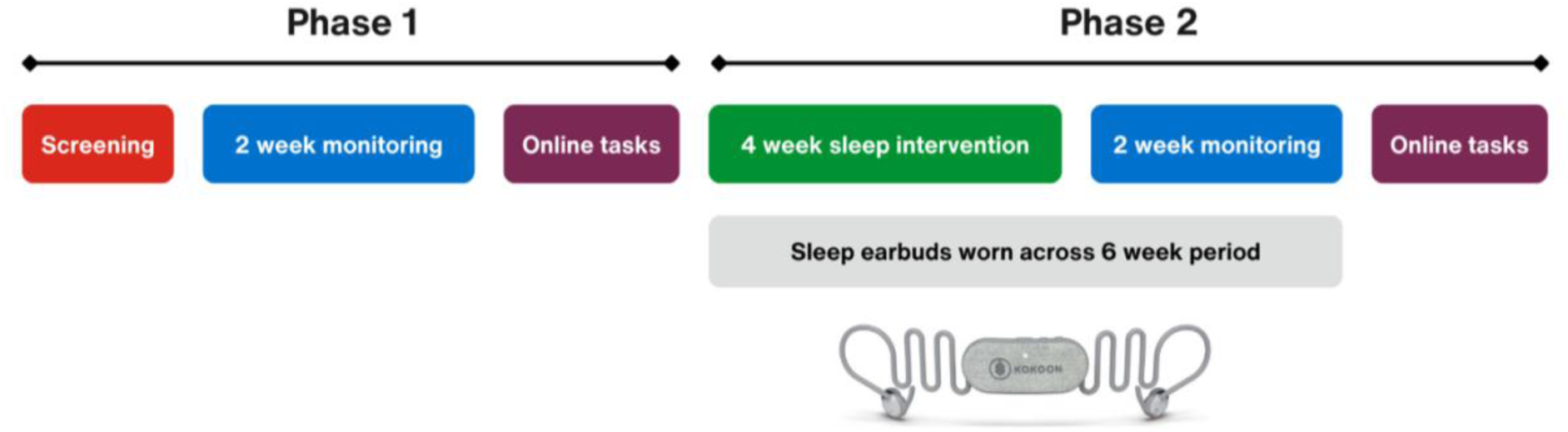
*EClocker Study* overview and timeline for Phase 1 and Phase 2

Sleep earbuds for *EClocker Phase 2* were worn for 6-weeks with a two-week monitoring period repeated from *EClocker Phase 1*. This involved identical study procedures with daily smartphone-based Experience Sampling Methods (ESM) and actigraphy (ActiGraph wGT3X-BT devices; ActiGraph LLC, Pensacola, USA) (*device data not reported here*). Momentary ESM and daily sleep and affective diaries were completed using the ExpiWell smartphone app (a commercial experience sampling platform; https://www.expiwell.com/).

Demographic and clinical characteristics of the nurses (N=27) at study entry are presented in **Table 1**. Nurses completed an online screening questionnaire (Qualtrics) at baseline during *EClocker Phase 1* to capture depressive symptoms (Patient Health Questionnaire; PHQ-9) [56–59], bipolar disorder (Mood Disorder Questionnaire; MDQ) [60], stress (Perceived Stress Scale; PSS-10) [61], sleep disorders (Global Sleep Assessment Questionnaire; GSAQ) [62], biological rhythm disruption (Biological Rhythms Interview of Assessment in Neuropsychiatry; BRIAN) [63] and chronotype (Morningness-Eveningness Questionnaire, MEQ; and The Caen Chronotype Questionnaire, CCQ) [64–68]. Additional demographic details, NHS employment history and NHS work shift patterns were also recorded. Most nurses identified as female (88.9%), were aged 26 to 43 years, and were of White ethnic origin (44.4%), Asian or Asian British (33.3%), or Black, Black British, Caribbean or African (18.5%). Shift information, number of nights, frequency of workdays, and average hours worked across Phase 1 and Phase 2 are summarised in **Table 2**.

**Table 1.**
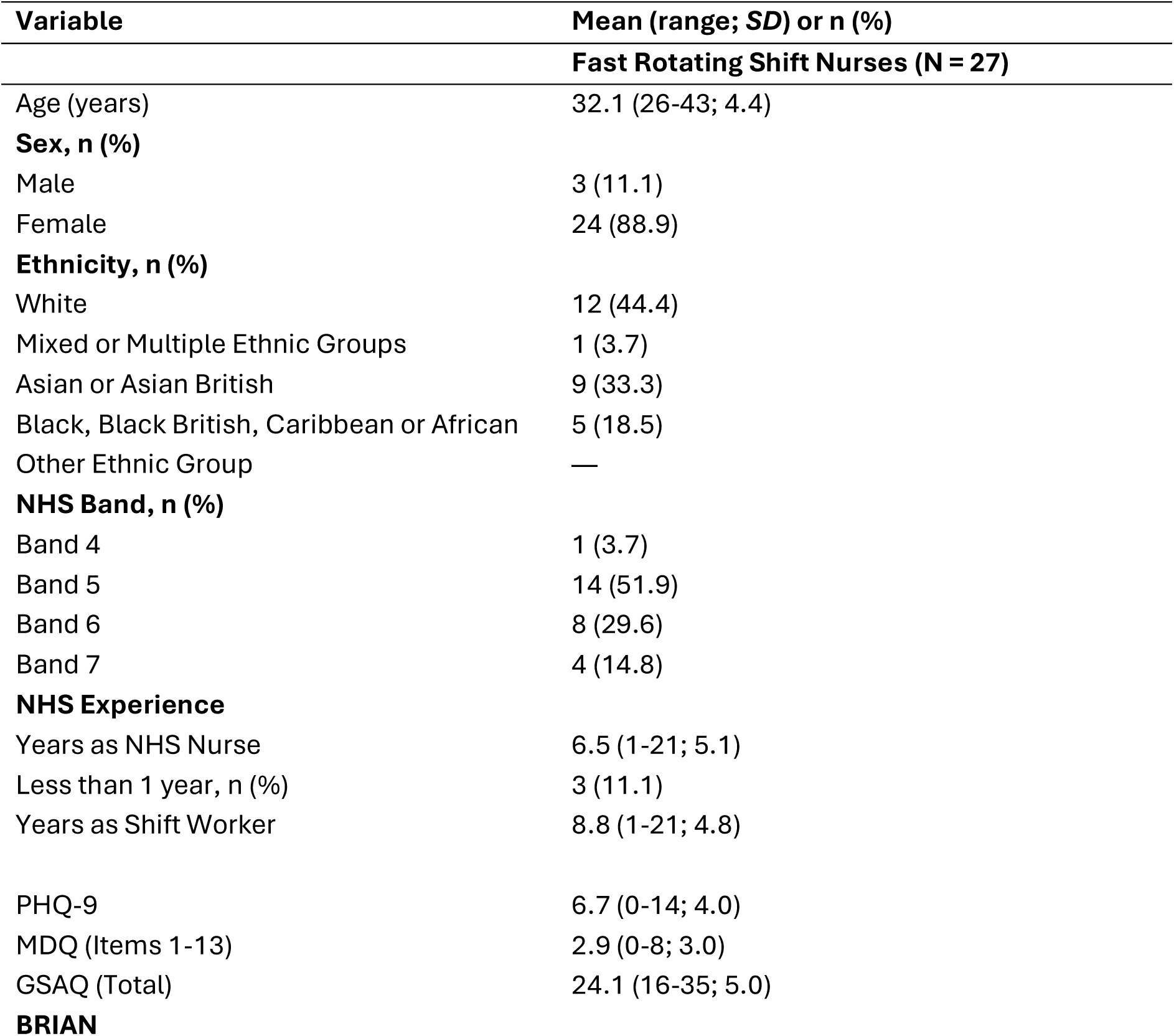
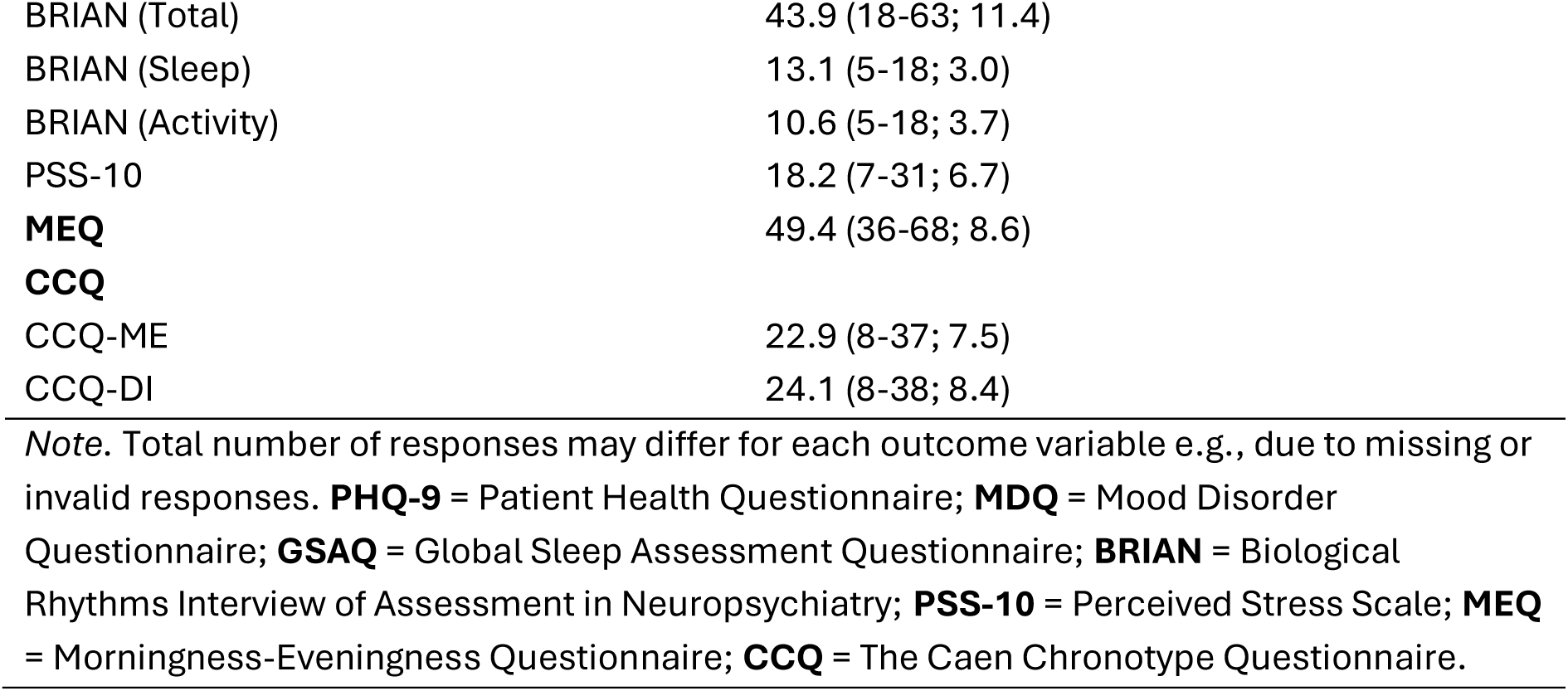
Sociodemographic and clinical characteristics of NHS nurses in the pilot sleep earbud intervention (N = 27). Variables were recorded at study entry (*EClocker Phase 1* screening responses)

| Variable | Mean (range; *SD*) or n (%) |
| --- | --- |
| **Fast Rotating Shift Nurses (N = 27)** |  |
| Age (years) | 32.1 (26-43; 4.4) |
| **Sex, n (%)** |  |
| Male | 3 (11.1) |
| Female | 24 (88.9) |
| **Ethnicity, n (%)** |  |
| White | 12 (44.4) |
| Mixed or Multiple Ethnic Groups | 1 (3.7) |
| Asian or Asian British | 9 (33.3) |
| Black, Black British, Caribbean or African | 5 (18.5) |
| Other Ethnic Group | — |
| **NHS Band, n (%)** |  |
| Band 4 | 1 (3.7) |
| Band 5 | 14 (51.9) |
| Band 6 | 8 (29.6) |
| Band 7 | 4 (14.8) |
| **NHS Experience** |  |
| Years as NHS Nurse | 6.5 (1-21; 5.1) |
| Less than 1 year, n (%) | 3 (11.1) |
| Years as Shift Worker | 8.8 (1-21; 4.8) |
| PHQ-9 | 6.7 (0-14; 4.0) |
| MDQ (Items 1-13) | 2.9 (0-8; 3.0) |
| GSAQ (Total) | 24.1 (16-35; 5.0) |
| **BRIAN** |  |
| BRIAN (Total) | 43.9 (18-63; 11.4) |
| BRIAN (Sleep) | 13.1 (5-18; 3.0) |
| BRIAN (Activity) | 10.6 (5-18; 3.7) |
| PSS-10 | 18.2 (7-31; 6.7) |
| **MEQ** | 49.4 (36-68; 8.6) |
| **CCQ** |  |
| CCQ-ME | 22.9 (8-37; 7.5) |
| CCQ-DI | 24.1 (8-38; 8.4) |

Note. Total number of responses may differ for each outcome variable e.g., due to missing or invalid responses. **PHQ-9** = Patient Health Questionnaire; **MDQ** = Mood Disorder Questionnaire; **GSAQ** = Global Sleep Assessment Questionnaire; **BRIAN** = Biological Rhythms Interview of Assessment in Neuropsychiatry; **PSS-10** = Perceived Stress Scale; **MEQ** = Morningness-Eveningness Questionnaire; **CCQ** = The Caen Chronotype Questionnaire.

**Table 2.**
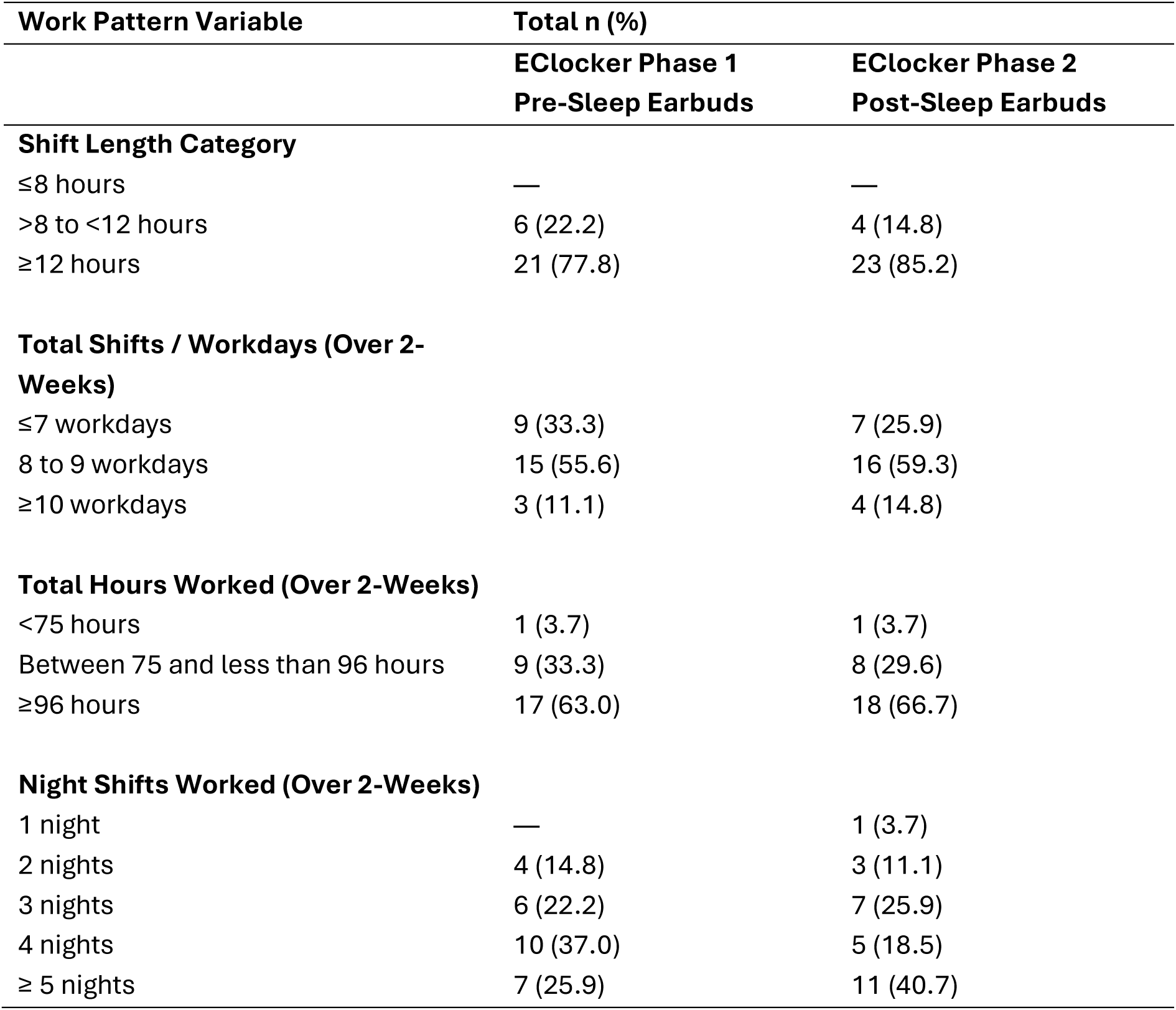
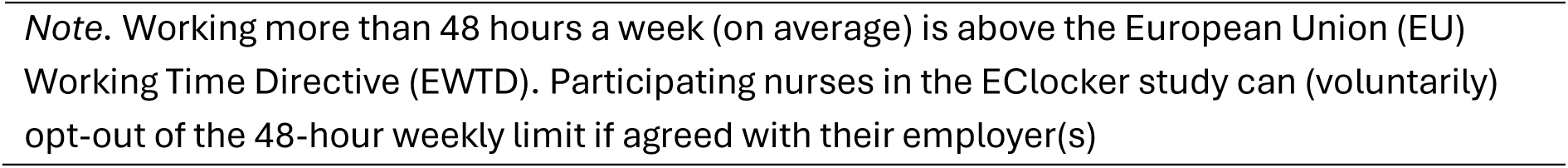
Average shift length, total number of workdays, night shifts, and overall hours worked (including overtime) for NHS fast rotating shift nurses (N = 27) who took part in *EClocker Phase 1* (pre-sleep earbuds) and *EClocker Phase 2* (post-sleep earbuds)

| Work Pattern Variable | Total n (%) |  |
| --- | --- | --- |
|  | EClocker Phase 1<br>Pre-Sleep Earbuds | EClocker Phase 2<br>Post-Sleep Earbuds |
| **Shift Length Category** |  |  |
| ≤8 hours | — | — |
| >8 to <12 hours | 6 (22.2) | 4 (14.8) |
| ≥12 hours | 21 (77.8) | 23 (85.2) |
| **Total Shifts / Workdays (Over 2-Weeks)** |  |  |
| ≤7 workdays | 9 (33.3) | 7 (25.9) |
| 8 to 9 workdays | 15 (55.6) | 16 (59.3) |
| ≥10 workdays | 3 (11.1) | 4 (14.8) |
| **Total Hours Worked (Over 2-Weeks)** |  |  |
| <75 hours | 1 (3.7) | 1 (3.7) |
| Between 75 and less than 96 hours | 9 (33.3) | 8 (29.6) |
| ≥96 hours | 17 (63.0) | 18 (66.7) |
| **Night Shifts Worked (Over 2-Weeks)** |  |  |
| 1 night | — | 1 (3.7) |
| 2 nights | 4 (14.8) | 3 (11.1) |
| 3 nights | 6 (22.2) | 7 (25.9) |
| 4 nights | 10 (37.0) | 5 (18.5) |
| ≥ 5 nights | 7 (25.9) | 11 (40.7) |

*Note.* Working more than 48 hours a week (on average) is above the European Union (EU) Working Time Directive (EWTD). Participating nurses in the EClocker study can (voluntarily) opt-out of the 48-hour weekly limit if agreed with their employer(s)

### Ethical Considerations

The *EClocker Study* ethics approval was provided by the Research Ethics Committee at King’s College London (HR-19/20-17792). The study was also approved by the Health Research Authority (HRA) and all participating NHS Trusts, Integrated Research Application System (IRAS) (289592). NHS organisations operated as Participant Identification Centres (PICs). All participants provided informed consent to participate in the study before taking part.

### Sleep Earbuds

The Kokoon NightBuds (**Figure S1 in Multimedia Appendix 1**) were developed by Kokoon Technology Ltd. Since completion of the *EClocker Phase 2* data collection, Kokoon partnered with Philips (Koninklijke Philips N.V.) and rebranded the Kokoon NightBuds in 2023 to Philips ‘Sleep headphones’ along with the Kokoon companion sleep app. The sleep earbuds and paired smartphone app (MyKokoon) used in *Phase 2* were therefore earlier, first-generation 2022 devices – Philips has since discontinued production of these sleep headphones (as of 2026). The company (Kokoon Technology Ltd.) was not involved with the *EClocker Study* at any phase, only to supply the consumer-grade devices as an industry partner and offer technical support to the research team if needed. Nurses were instructed to wear the sleep earbuds in both ears during their major sleep episodes, nap periods (including prophylactic naps), or any attempted sleep over the 6-week study period. Main sleep episodes were either at night or during the day (e.g., after a night shift had ended).

The sleep earbuds were paired with the ‘MyKokoon’ smartphone app which was downloaded on participants’ personal devices – see **Figure S2** in **Multimedia Appendix 1** for an example of the app interface. The research team provided secure log-in details to access the paired MyKokoon app to ensure confidentiality and anonymity. Sensor data from the earbuds were integrated into the MyKokoon app which provided daily sleep summaries and metrics for the user. The smartphone app contained an audio library that delivered personalised relaxation exercises (e.g., meditation, deep breathing, progressive muscle relaxation, sleep visualisation) and sleep techniques drawn from cognitive behavioural therapy for insomnia (CBT-I). The earbuds were designed to mask ambient sounds and external noise disturbances to promote sleep episodes and aid relaxation. Sleep metrics recorded by the earbuds included: number of sleep sessions; total sleep time (TST); sleep-wake timings; sleep onset latency (SOL); number of sleep interruptions; wakefulness periods; sleep efficiency (SE); and sleep staging (awake, light, deep, REM and total epochs).

The sleep earbuds used a Photoplethysmogram (PPG) sensor to measure heart rate and heart rate variability. Sleep tracking technology was based on the in-ear PPG optical heart rate sensor. The device used 5.4 mm silicone earbuds with an overall battery life of ≥10 hours. Biometric sensors embedded for sleep monitoring were comparable with other commercially available in-ear products at the time of data collection (e.g., Bose Sleepbuds, Ozlo Sleepbuds, Amazfit ZenBuds). Sensor detection included adaptive audio auto-fade and noise-masking (e.g., the device faded audio from the app, then introduced either coloured noise to mask disturbances or switch off the device).

### Primary Outcomes

Acceptability, helpfulness, and perceived effectiveness of the pilot sleep earbud intervention was evaluated through a brief feedback questionnaire (Qualtrics) at the end of the study. Questionnaire items were in line with prior sleep aid studies for noise-masking or multi-audio interventions (e.g., using white and pink ‘noise’ or music) [52, 69–71], sleep trials such as Sleepio [72], and smartphone-based sleep interventions [73].

The degree to which nurses found the sleep earbuds effective, helpful, and acceptable were evaluated with adapted items from the Treatment Acceptability Questionnaire (6- item TAQ) [74]. The TAQ has previously been used to evaluate sleep treatment acceptability such as digital CBT-I interventions for insomnia [75]. All three adapted TAQ items were rated on a 7-point scale (e.g., responses ranged from 1 = Very ineffective to 7 = Very effective; 1 = Very unhelpful to 7 = Very helpful; 1 = Very unacceptable to 7 = Very acceptable). A qualitative question adapted from Smejka, Henry [72] also explored individual experiences of using the sleep earbuds (‘*Overall, how did you find using the Kokoon Sleep Headphones? Have you noticed a change?’*).

Sleep outcome items (**Table 3**) were adapted from the PROMIS Sleep Disturbance (Short Form 8a; Patient-Reported Outcomes Information System) [76] and Falkenberg, Tungland [77]. Nurses rated the impact of the sleep earbuds on their sleep onset latency (SOL), sleep quality (SQ), sleep duration (TST) and sleep disturbance. Each item was rated on a 7-point Likert scale from 1 (Strongly disagree) to 7 (Strongly agree).

**Table 3.** Acceptability, helpfulness, and perceived effectiveness after 6-weeks use of the sleep earbuds from *EClocker Phase 2* fast rotating shift nurses (N = 27)

| Statements | Mean (SD) | Median (IQR) | Min | Max |
| --- | --- | --- | --- | --- |
| **Sleep Outcomes** |  |  |  |  |
| I fell asleep more easily | 4.4 (2.1) | 5.0 (5) | 1 | 7 |
| My sleep quality improved | 4.1 (1.8) | 4.0 (3) | 1 | 7 |
| I slept for longer | 3.4 (1.6) | 3.0 (3) | 1 | 7 |
| My sleep was less disturbed | 3.7 (1.7) | 3.0 (3) | 1 | 7 |
| **Acceptability Measures** |  |  |  |  |
| Effectiveness | 4.0 (1.6) | 4.0 (2) | 1 | 7 |
| Helpfulness | 4.2 (1.9) | 5.0 (3) | 1 | 7 |
| Acceptability | 4.1 (1.7) | 4.0 (3) | 1 | 7 |

### Secondary Outcomes

Mean (SD) and Median (IQR) scores for secondary outcome measures are summarised in **Table 4**. These measures included the Pittsburgh Sleep Quality Index (PSQI) [78] to assess sleep disturbance and sleep quality, Maslach Burnout Inventory – Human Services Survey for Medical Personnel (MBI-HSS MP) [79] to evaluate burnout symptoms, the Positive and Negative Affect Schedule (PANAS-GEN) [80] to assess affect, Difficulties in Emotion Regulation Scale Short Form (DERS-SF) [81, 82] to assess emotion dysregulation and the Emotion Reactivity Scale (ERS) [83] to assess trait emotional reactivity. These secondary outcome measures (PSQI, MBI-HSS MP, PANAS-GEN, DERS-SF and ERS) were completed across two time points (pre- and post-sleep earbud intervention). A recent meta-analysis [84] reported how improving sleep (as a transdiagnostic target) also improved mental health outcomes, stress, and symptoms of depression and anxiety. Sleep improvements from the pilot earbud intervention may have cumulative and restorative effects on secondary outcome symptoms in reducing burnout and yield improved affective states and emotional reactivity.

**Table 4.** Pre-and post-sleep earbud intervention scores for secondary outcome measures (Fast Rotating Shift Nurses, N = 27)

| Outcome Variable | EClocker Phase 1<br>Pre-Sleep Earbuds |  | EClocker Phase 2<br>Post-Sleep Earbuds |  | Median of<br>Differences |
| --- | --- | --- | --- | --- | --- |
|  | Mean (SD) | Median<br>(IQR) | Mean (SD) | Median<br>(IQR) | Wilcoxon<br>Signed Ranks<br>Test, *p* |
| Sleep Quality (PSQI Global) | 7.9 (3.5) | 7.0 (3) | 7.0 (2.1) | 7.0 (3.0) | .114 |
| **Burnout (MBI-HSS MP)** |  |  |  |  |  |
| Emotional Exhaustion | 27.9 (11.7) | 29.0 (19) | 30.4 (11.8) | 33.0 (19) | .156 |
| Depersonalization | 8.9 (5.0) | 8.0 (8) | 10.5 (5.3) | 10.0 (8) | .065 |
| Personal<br>Accomplishment | 33.3 (6.0) | 34.0 (6) | 32.9 (7.3) | 34.0 (8) | .638 |
| **Affective State (PANAS-**<br>**GEN)** |  |  |  |  |  |
| Positive Affect (PA) | 33.7 (7.3) | 34.0 (12) | 34.9 (5.9) | 36.0 (11) | .286 |
| Negative Affect (NA) | 18.0 (5.5) | 18.0 (10) | 19.0 (4.8) | 20.0 (8) | .265 |
| Emotion Regulation<br>(DERS-SF Total) | 38.0 (11.7) | 36.0 (17) | 39.6 (11.3) | 37.0 (17) | .658 |
| Emotion Reactivity (ERS<br>Total) | 27.7 (17.1) | 24.0 (19) | 27.2 (14.6) | 26.0 (18) | .722 |
| *Note.* Total number of responses may differ for each outcome variable e.g., due to missing or invalid responses. **PSQI** = Pittsburgh Sleep Quality Index; **MBI-HSS MP** = Maslach Burnout Inventory – Human Services Survey for Medical Personnel; **PANAS-GEN** = Positive and Negative Affect Schedule; **DERS-SF** = Difficulties in Emotion Regulation Scale Short Form; **ERS** = Emotion Reactivity Scale. |  |  |  |  |  |

Sleep earbuds were worn for six-weeks during the intervention period. Nurses also completed concurrent daily subjective (Consensus Sleep Diary; CSD) [85] and objective (actigraphy; wGT3X-BT) sleep measures and smartphone-based ESM monitoring for the final two-weeks of the intervention. These measures were identical to the two-week monitoring period in *EClocker Phase 1*. Day-to-day outcome measures are summarised in **Table 5** for nightly sleep (TST, SOL, SE, SQ), mood, and affect (PA / NA) reported for *EClocker Phase 1* (pre-earbud intervention) and *EClocker Phase 2* (post-earbud intervention) and were based on previous reviewed literature [86]. Histogram distributions for these daily smartphone-based ESM variables are presented in **Figure 5**.

**Figure 2.**
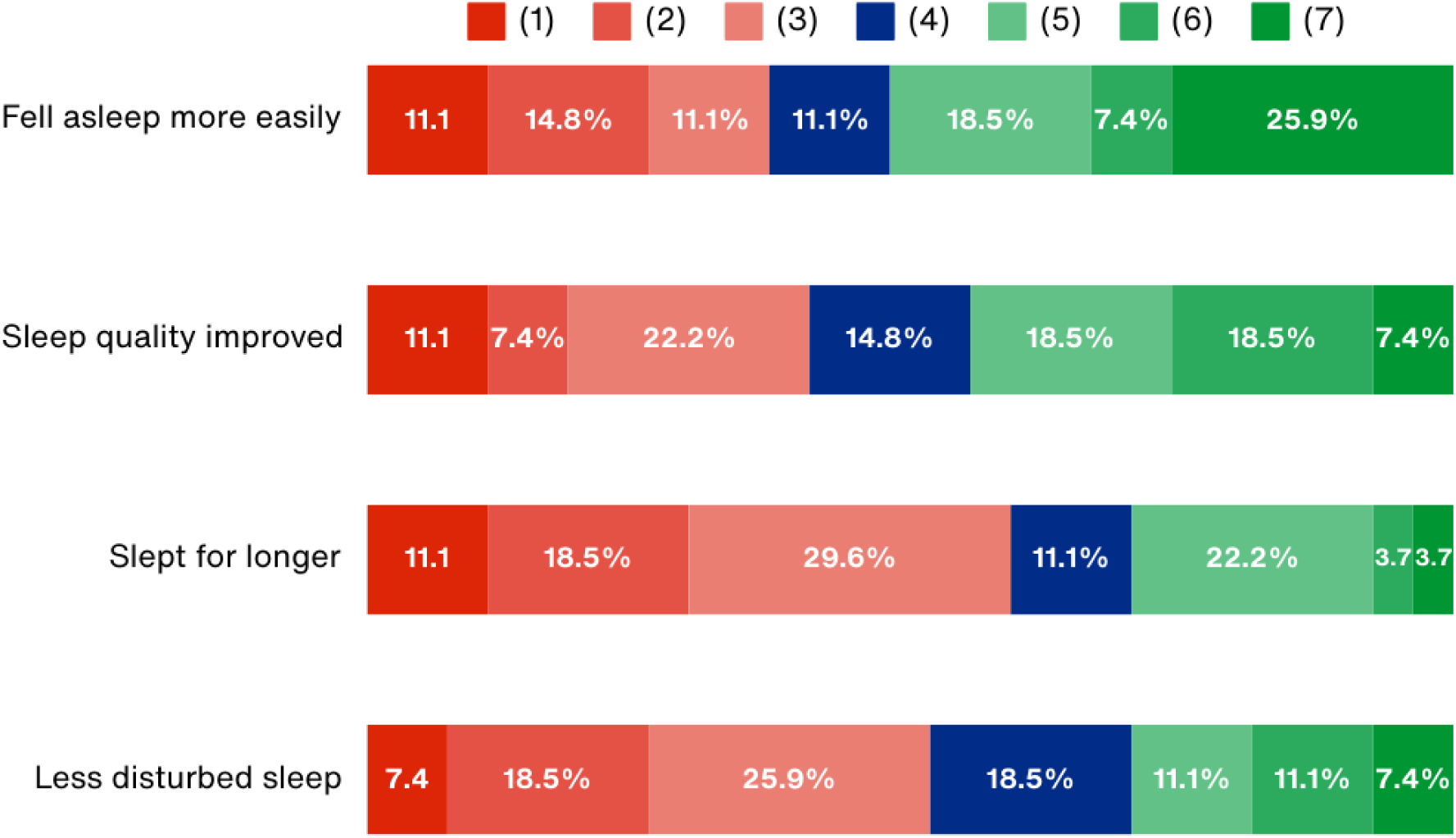
NHS nurses rated the impact of the sleep earbuds on their sleep onset latency (SOL), sleep quality (SQ), sleep duration (TST) and sleep disturbance after 6-weeks. Each question was rated on a 7-point Likert scale from 1 (Strongly disagree), 2 (Disagree), 3 (Somewhat disagree), 4 (Neither agree nor disagree), 5 (Somewhat agree), 6 (Agree) to 7 (Strongly agree)

**Figure 3.**
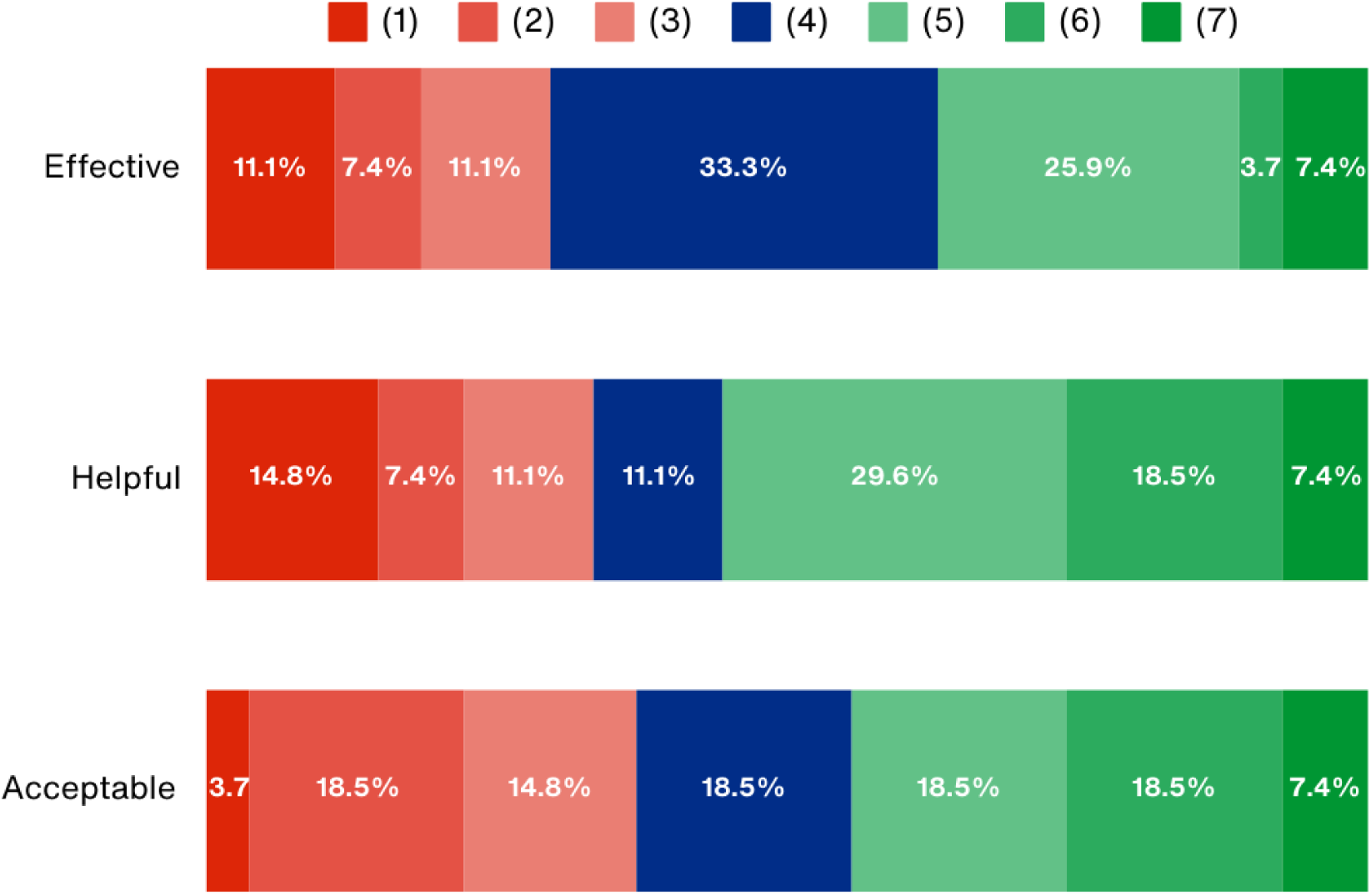
NHS nurses rated the perceived level of effectiveness, helpfulness, and acceptability of the sleep earbuds after 6-weeks. Adapted TAQ items were rated on a 7- point scale (e.g., responses ranged from 1 = Very ineffective to 7 = Very effective; 1 = Very unhelpful to 7 = Very helpful; 1 = Very unacceptable to 7 = Very acceptable)

**Figure 4.**
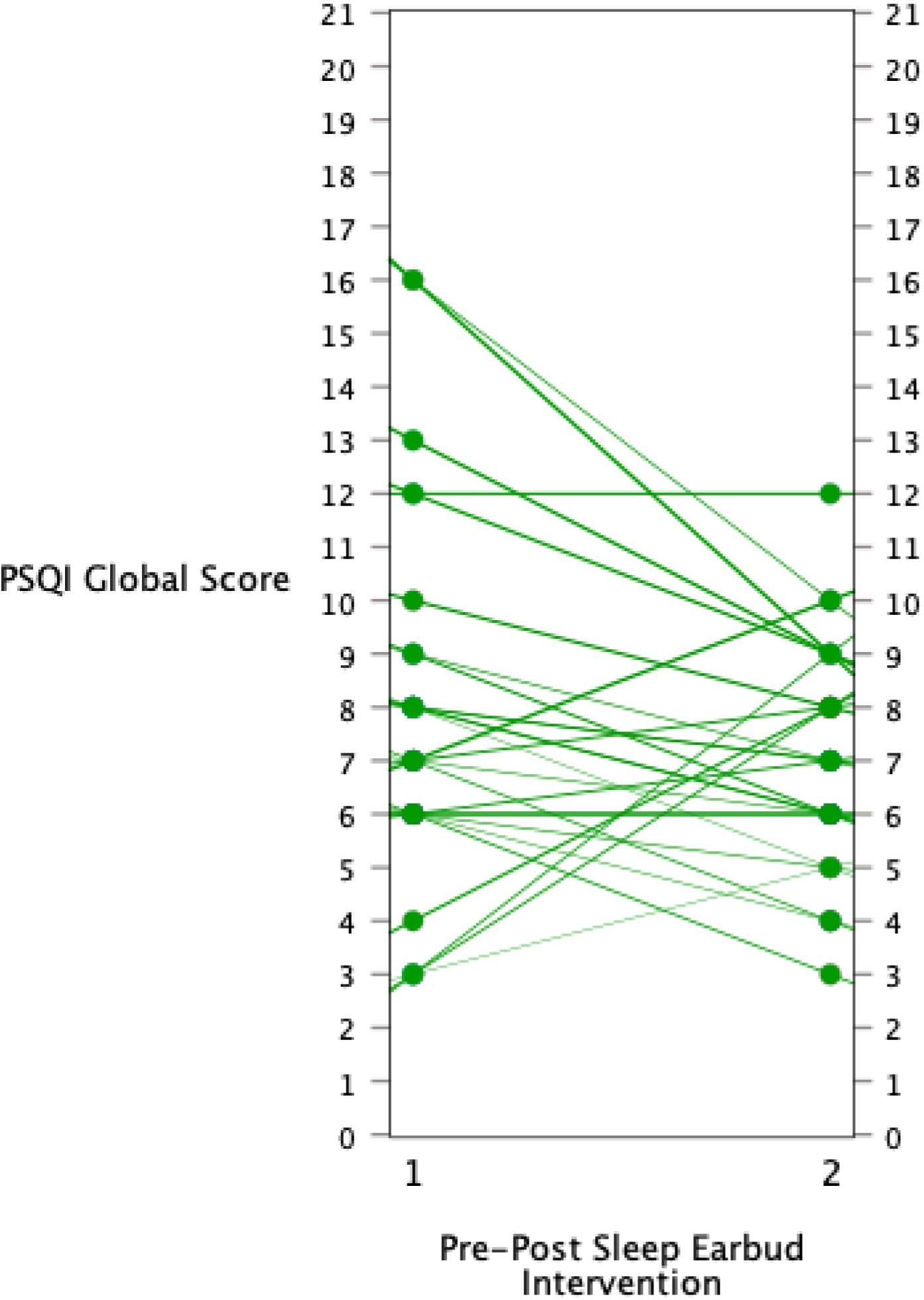
Spaghetti plot visualising sleep quality trajectories (PSQI Global) from pre-to- post sleep earbud intervention. Each line (green) represents an individual NHS Fast Rotating Shift Nurse (N = 27). Negative gradients denote improved sleep scores

**Figure 5.**
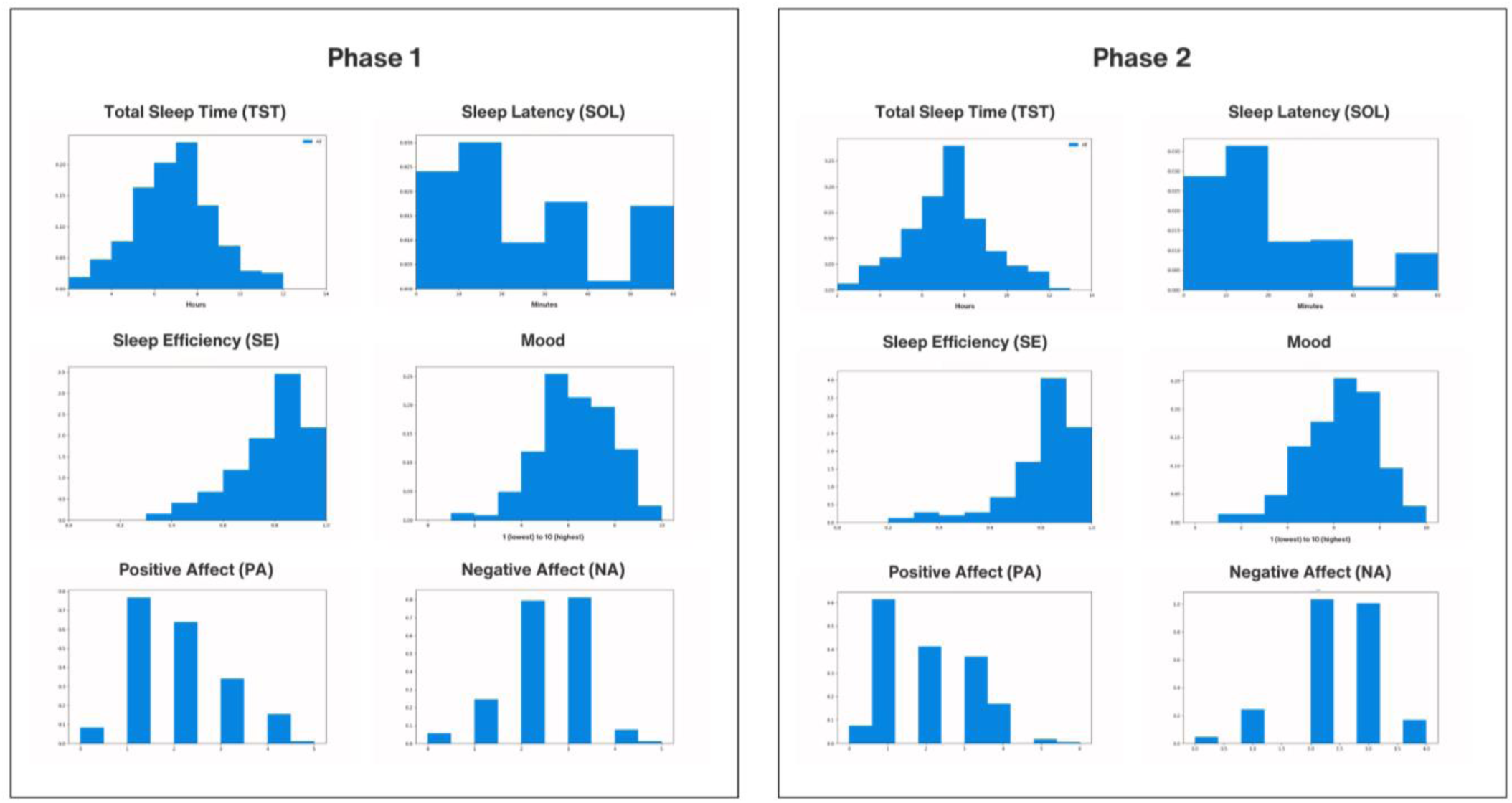
Histogram distributions for daily smartphone-based ESM variables (averaged over the 2-week data collection) for *EClocker Phase 1* (left) and *EClocker Phase 2* (right). Sleep (TST, SOL, SE), mood (rated low to high), and affect (PA and NA) outcomes are shown for all NHS fast rotating shift nurses (N = 27) who participated in Phase 1 and Phase 2

**Table 5.** Smartphone-based ESM variables self-reported by NHS fast rotating shift nurses (N = 27) who took part in *EClocker Phase 1* and *EClocker Phase 2*. Sleep, mood, and affect scores are averaged over the 2-week data collection for Phase 1 and Phase 2

| Variables | EClocker Phase 1<br>Pre-Sleep Earbuds |  |  |  | EClocker Phase 2<br>Post-Sleep Earbuds |  |  |  |
| --- | --- | --- | --- | --- | --- | --- | --- | --- |
|  | Mean | SD | Min | Max | Mean | SD | Min | Max |
| Total Sleep Time (TST) | 6.88 | 1.88 | 2.50 | 11.88 | 7.18 | 1.94 | 2.00 | 12.33 |
| Sleep Latency (SOL) | 31.52 | 38.63 | 0.00 | 325.0 | 20.87 | 24.87 | 0.00 | 180.0 |
| Sleep Efficiency (SE) | 0.79 | 0.14 | 0.38 | 1.00 | 0.81 | 0.14 | 0.27 | 1.00 |
| Sleep Quality (SQ) | 3.19 | 0.72 | 2.00 | 4.00 | 3.27 | 0.73 | 2.00 | 4.00 |
| Mood | 6.02 | 1.60 | 1.00 | 10.00 | 6.05 | 1.60 | 1.00 | 10.00 |
| Positive Affect (PA) | 1.88 | 1.04 | 0.00 | 5.00 | 2.02 | 1.16 | 0.00 | 6.00 |
| Negative Affect (NA) | 2.32 | 0.87 | 0.00 | 5.00 | 2.40 | 0.83 | 0.00 | 4.00 |

*Note.* Sleep parameters are from the daily Consensus Sleep Diary (CSD); **TST** in hours, **SOL** in minutes, **SQ** scored on a 1-5 scale; **Mood** scored on a 1-10 scale and averaged daily; **Affect** scored on a 0-7 scale and averaged daily for PA and NA

### Statistical Analysis

Quantitative analyses were undertaken in IBM SPSS Statistics (V.29) for descriptive summaries of baseline screening characteristics (**Table 1**), work shift patterns (**Table 2**), pre-and post-sleep earbud intervention scores (**Table 4**) including Wilcoxon Signed Ranks Tests and Spaghetti plots (**Figure 4**). Quantitative analyses were undertaken in Python for smartphone-based ESM variables (**Table 5**) and histogram distributions (**Figure 5**).

#### Power

A power analysis calculated a priori during *EClocker* protocol study design had estimated an adequate sample of 66 fast rotating shift nurses, with a medium effect size (d=0.50), 95% power, and a stringent alpha level (α = 0.01). Sample size, however, was limited for *EClocker Phase 2* by pragmatic challenges such as the Covid-19 pandemic, study timeframe, device availability and lab resources.

## Results

### Primary Outcomes

Nurses subjectively rated whether the earbuds improved their sleep after 6-weeks of use (**Figure 2**) and the overall acceptability, helpfulness, and perceived effectiveness of the earbuds (**Figure 3**). Descriptive scores for primary outcomes are presented in (**Table 3**) and secondary outcomes in (**Table 4**) and (**Table 5**). Median (IQR) values are provided (in addition to Mean and Minimum-Maximum values) due to non-normal distributions of certain measure items.

### Secondary Outcomes

#### Descriptive Results

Following six-weeks use of the sleep earbuds, 63% of NHS fast rotating shift nurses (N=27) reported a decrease in sleep disturbance scores (PSQI Global). A third of nurses (33.3%) subjectively perceived improved sleep latency (SOL), sleep efficiency (SE), and better daytime dysfunction scores (PSQI subcomponents). Around a quarter of nurses (25.9%) rated improved sleep quality (SQ) and a fifth (22.2%) reported higher sleep duration (TST) (PSQI subcomponent scores). Improvements in global sleep quality symptoms are visually presented in **Figure 4** on an individual basis. No concomitant changes emerged for secondary outcome measures (**Table 4**) of burnout (MBI-HSS MP), affective state (PANAS-GEN), emotion regulation (DERS-SF) or emotion reactivity (ERS).

Sleep duration (TST) and latency (SOL) which was reported daily (CSD sleep diaries) over two-weeks in *EClocker Phase 1* and *Phase 2* had improving trends post-sleep earbud intervention. Nurses (on average) reported sleeping longer and falling asleep faster, with total sleep (TST) increasing by 18 minutes and average sleep onset latency (SOL) decreasing by almost 11 minutes (difference in average TST and SOL from pre-to-post across the sample; **Table 5**).

Since the primary objectives of *EClocker Phase 2* were to evaluate acceptability and initial implementation of the sleep earbuds in a shift working population, *p*-values for Wilcoxon Signed Ranks Tests (**Table 4**) are reported for descriptive (rather than inferential) purposes; this is in line with descriptive reporting recommendations for non-randomised pilot studies [87, 88]. Therefore, whilst almost two thirds (63%) of NHS fast rotating shift nurses had improved sleep quality scores post-sleep earbud use (PSQI Global), the relative magnitude and clinically meaningful effect size estimates of the pilot intervention are not interpretable [89].

## Discussion

### Principal Findings

A pilot noise-masking earbud intervention showed promising (albeit small) sleep improvement trends after six-weeks in a sample of NHS fast rotating shift nurses. To our knowledge, this is the first study to pilot noise-masking earbuds as a sleep aid for a cohort of shift workers in Europe or the UK. Sleep earbuds may have the potential to be a relatively cost-effective, accessible, and minimally time-intensive digital intervention for shift workers. Clinical significance and the magnitude of sleep improvement change, however, was not evaluated in this small, preliminary study.

Almost two thirds of nurses (63%) subjectively reported (small) reductions in general sleep disturbance symptoms post-sleep earbud use (PSQI Global scores). One in four nurses experienced perceived sleep quality improvements (SQ; 25.9%), one in five subjectively slept longer (TST; 22.2%), and a third reported falling asleep faster (SOL; 33.3%), better sleep efficiency (SE; 33.3%) and improved daytime dysfunction (33.3%) (PSQI subcomponent scores). Sleep diaries (CSD) collected daily also demonstrate small improvements post-sleep earbud use; nurses slept an average 18 minutes longer (TST) and fell asleep an average 11 minutes faster (SOL). Although sleep duration changes may not be clinically meaningful, average TST after the intervention (7 hours 11 minutes) met consensus sleep expert recommendations of ≥7 hours for adults aged 18- 60 years [90, 91].

Retention rate at six-weeks was 82%, with a dropout (18%) consistent with prior Randomised Controlled Trial (RCT) sleep interventions with nursing staff [5]. Sleep earbuds were reported as somewhat to very helpful (56%) by over half of NHS fast rotating shift nurses; 44% rated the earbuds as somewhat to very acceptable; and over a third (37%) reported the earbuds as somewhat to very effective. Around half of nurses (52%) agreed (somewhat to strongly) using the earbuds improved their sleep latency (SOL; fell asleep more easily); 44% agreed (somewhat to strongly) their sleep quality (SQ) improved; and almost a third (30%) agreed (somewhat to strongly) they slept for longer (TST) and had less disturbed sleep. No severe adverse events were reported by nurses, and the earbuds were generally well tolerated, aside from mild in-ear discomfort. Pharmacological treatments, by contrast, such as wake-promoting agents used for shift work disorder (e.g., modafinil and armodafinil) have more severe medication side effects such as headaches, nausea, nervousness and nasopharyngitis [92, 93].

These preliminary findings indicate consumer-grade sleep earbuds may be beneficial for some (but not all) night shift workers, consistent with prior interventions in the United States using the Bose SleepBuds [53, 54]. As with other sleep interventions, this is not a “one-size fits all” approach and barriers for larger uptake may be attributed to baseline shift worker preferences. Recent evidence from Acevedo, Mu [1], for example, demonstrate the relevance of individual factors and personal preferences in nurses’ endorsement of sleep interventions. Acceptability of interventions for shift workers, however, remains an under-studied area [14, 94]. Acceptability and satisfactory ratings in this study, appear to be impacted by nurses’ reports of residual pain, in-ear pressure, or uncomfortable fit following prolonged use of earbuds at night. These wearability barriers - cited among qualitative responses - have similarly been reported by healthcare staff using Bose SleepBuds [54]. Perceived baseline earbud comfort, ergonomic fit ratings, and initial satisfaction could therefore contribute to poorer sleep intervention outcomes, adherence, and discontinuation rates for some nurses [95–97]. Pre-treatment acceptability ratings, for example, predict therapy adherence, session attendance, drop-out rates and sleep treatment response among patients with insomnia [98, 99].

### Future Directions

This was an uncontrolled pilot study with a modest sample of (self-selecting) fast rotating shift nurses. Results should be interpreted with caution as there was not sufficient analytical power to evaluate the magnitude of intervention effect sizes. Accordingly, a larger controlled trial study is needed with an extended intervention period beyond six-weeks, given sleep improvements may emerge at a later timepoint with consistent earbud use. Follow-up assessments are also required to establish immediate to long-term sleep benefits and longevity of behaviour change. Pre- to post-sleep changes were small and may not be of clinical interest; this may reflect nurses’ longer-term interactions and challenges with shiftwork conditions, exposure to irregular schedules, circadian misalignment or sleep displacement. Indeed, 89% (N=24) of nurses taking part in the intervention had problematic baseline sleep difficulties indicative of potential shift work sleep disorder (GSAQ Item 4) and insomnia (GSAQ Item 1). Furthermore, NHS nurses also had roster changes from Phase 1 (pre-sleep earbuds) to Phase 2 (post-sleep earbuds) (**Table 2**). Average shift length and number of night shifts experienced was higher during Phase 2 (post-sleep earbuds), with more NHS nurses (14.8% increase) working ≥5-night shifts.

*EClocker Phase 2* was designed as a non-pharmacological ‘as required’ pilot intervention for which nurses could use the sleep earbuds at their own discretion. As this was the first pilot stage in evaluating initial acceptability, future work should compare the earbuds with similar commercially available in-ear devices (e.g., Ozlo Sleepbuds, Amazfit ZenBuds) or alternative widespread white noise sleep aids (see Riedy, Smith [52] for a recent review). Sleep earbud efficacy could also be evaluated with other healthcare populations (e.g., doctors or emergency service workers), individuals with a clinical diagnosis of shiftwork associated sleep disorder (SWD) or shift workers in other industry sectors. Noise-masking earbuds are commercially available and so have the potential to be widely disseminable, transdiagnostic, and scalable to the general population. Moreover, according to a report commissioned by the Royal Society for Public Health (RSPH), poor sleep is ubiquitous and the second most common health complaint in the UK, with around 1 in 5 individuals regularly sleeping poorly at night [100]. A fifth of young adults may also have a sleep disorder which warrants clinical review and treatment [101]. An accessible and relatively low-cost sleep intervention could therefore have significant long-lasting and public health benefits.

Sleep beliefs and knowledge held by nurses could be reported at baseline in future work. Prior studies have shown unhelpful, dysfunctional, or maladaptive sleep beliefs or myths can adversely impact sleep behaviours, influence appraisal of sleep, or the reporting of sleep-related difficulties [102–104]. Sleep aid use at baseline (e.g., use of blackout curtains, eye masks, earplugs) could also be assessed as per previous studies [54]. Adherence levels for the sleep earbuds including day-to-day usage, continued wear-time, and six-week intervention engagement will require second phase analyses to determine feasibility. Acceptability of the accompanying earbud app (MyKokoon in **Multimedia Appendix 1**) could be explored further with specific user-experience ratings. Additional research is needed to evaluate the audio type, specific noise characteristics, and length of time audio content was accessed, in relation to sleep changes. Instruments which monitor environmental noise levels could also be integrated in future studies (e.g., noise dosimeters). Post-night shift healthcare staff are at particular risk for ambient noise-induced sleep disturbance during the daytime [105–107]. Noise-masking earbuds could conceivably protect shift workers by filtering or blocking harmful exposure to external background environmental noise during daytime sleep episodes. Fewer noise-induced and auditory disruptions could promote more restorative and deep sleep, reduce sleep fragmentation, decrease micro-awakenings or arousals and aid sleep onset in shift workers [52, 105].

## Conclusions

Noise-masking earbuds may be a plausible, adjunctive intervention approach to improve sleep health for healthcare workers. There are no “one-size fits-all” approaches to improve sleep, however, and countermeasures or prevention strategies should be tailored for each shift worker (including underlying individual sleep-circadian phenotype) to ensure optimal adherence and treatment gain. Larger controlled studies with follow-up periods are required to establish long-term sleep earbud benefits.

## Supporting information

Multimedia Appendix 1

## Data Availability

All data produced in the present study are available upon reasonable request to the authors

## Acknowledgements

The authors thank the research participants, NHS staff, and NHS Foundation Trusts who contributed their time and efforts towards this research. The authors also thank Kokoon Technology Ltd. for providing the sleep earbuds for the purposes of this study.

## Contributors

Conceptualisation (RH, TCD, SS); Methodology (RH, TCD, SS); Data collection (RH); Formal analysis (RH, DWJ, NG); Writing - original draft preparation (RH); Writing - review and editing (all authors); Supervision (TCD, SS).

## Author Declaration

This article incorporates preliminary material revised and reworked from Doctoral thesis work [submitted by author Robert Hickman for the Doctor of Philosophy at King’s College London].

This paper represents independent research [part] funded by the National Institute for Health and Care Research (NIHR) Maudsley Biomedical Research Centre at South London and Maudsley NHS Foundation Trust and King’s College London. The views expressed are those of the author(s) and not necessarily those of the NIHR or the Department of Health and Social Care.

This work is partly supported by the Wellcome Trust [CHiP-D Study: Grant Number 227099/Z/23/Z] for authors RH, DJ, SS, TD.

## Data Availability

Data are available upon reasonable request to the corresponding author.

## Ethical Approval

This study involves human participants and was approved by the Research Ethics Committee at King’s College London (HR-19/20-17792). The study was also approved by the Health Research Authority (HRA) and all participating NHS Trusts, Integrated Research Application System (IRAS) (289592). NHS organisations operated as Participant Identification Centres (PICs). Participants gave informed consent to participate in the study before taking part.

**Abbreviations**
BRIAN: Biological Rhythms Interview Of Assessment In Neuropsychiatry
CBT-I: Cognitive Behavioural Therapy For Insomnia
CCQ: The Caen Chronotype Questionnaire
CSD: Consensus Sleep Diary
DERS-SF: Difficulties In Emotion Regulation Scale Short Form
ERS: Emotion Reactivity Scale
ESM: Experience Sampling Method
EWTD: European Union Working Time Directive
GSAQ: Global Sleep Assessment Questionnaire
IRAS: Integrated Research Application System
MBI-HSS MP: Maslach Burnout Inventory – Human Services Survey For Medical Personnel
MDQ: Mood Disorder Questionnaire
MEQ: Morningness-Eveningness Questionnaire
NHS: National Health Service
NICE: National Institute For Health And Care Excellence
NMC: Nursing And Midwifery Council
PANAS-GEN: Positive And Negative Affect Schedule
PHQ-9: Patient Health Questionnaire
PIC: Participant Identification Centre
PSQI: Pittsburgh Sleep Quality Index
PSS-10: Perceived Stress Scale
REM: Rapid Eye Movement
SE: Sleep Efficiency
SOL: Sleep Onset Latency
SQ: Sleep Quality
SWD: Shift Work Disorder
TST: Total Sleep Time

## Multimedia Appendix

**Multimedia Appendix 1:** Kokoon NightBuds and MyKokoon smartphone app user interface

