## Supplementary material for "Sleep Intervention for NHS Healthcare Shift Workers: A Pilot Study of Noise-Masking Earbuds": Multimedia Appendix 1

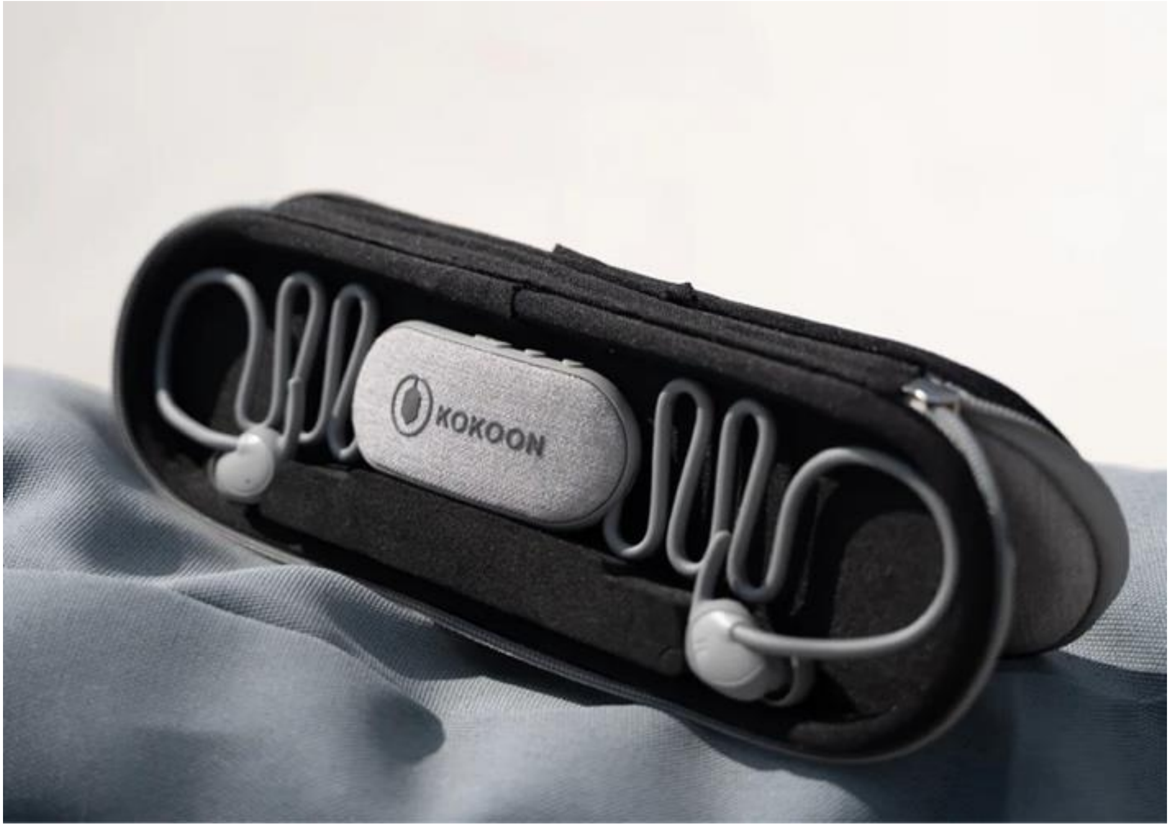


**Figure S1.** Kokoon NightBuds first-generation 2022 device (Kokoon Technology Ltd.) were worn by NHS nurses for 6-weeks during *EClocker Phase 2*

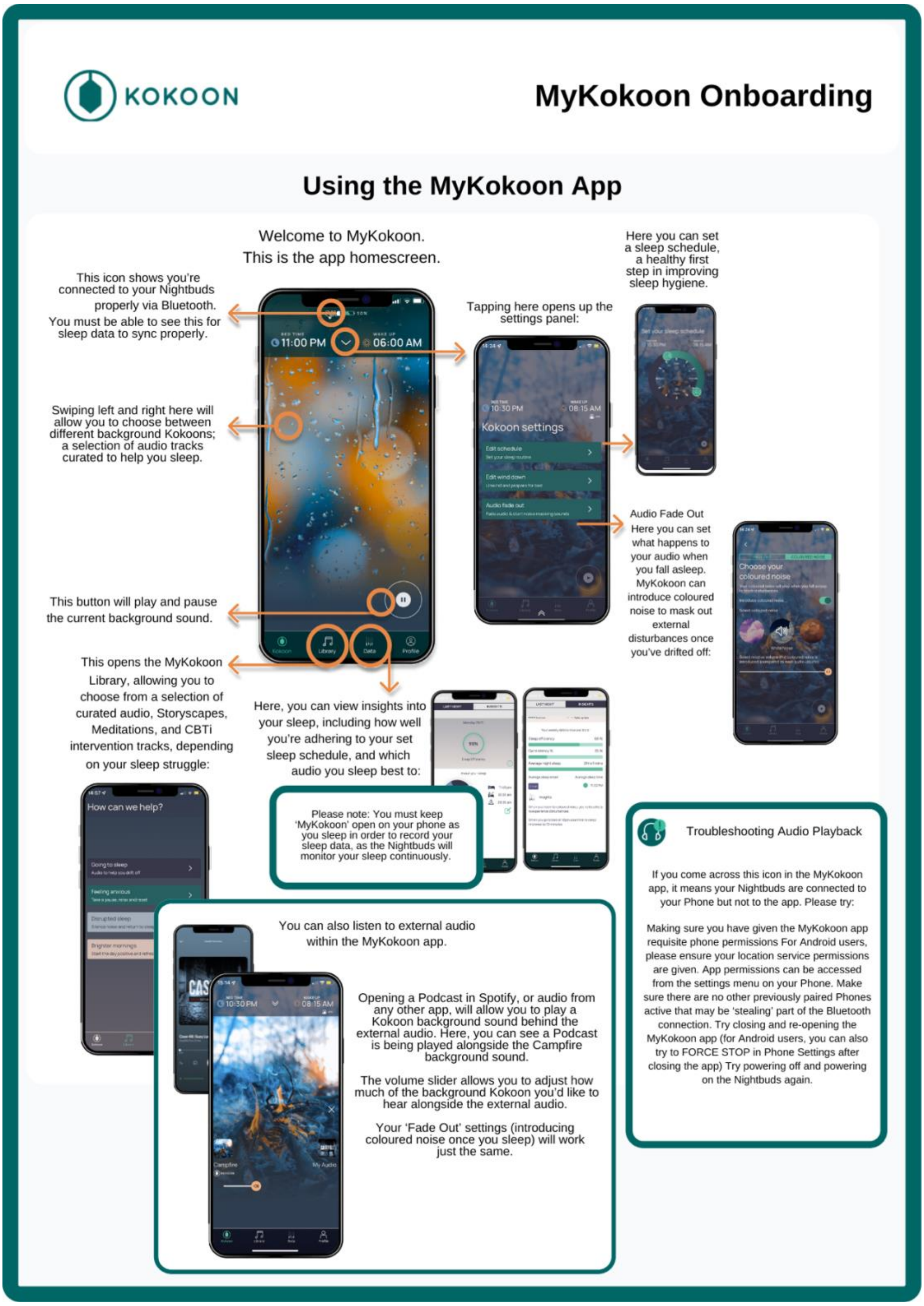


**Figure S2.** MyKokoon smartphone app user interface and onboarding instructions. App interface (2022) was an earlier version compatible with the first-generation Kokoon NightBuds (Kokoon Technology Ltd.)
